# Reported COVID-19 Incidence in Wisconsin High School Athletes During Fall 2020

**DOI:** 10.1101/2021.02.18.21251986

**Authors:** Phillip Sasser, Timothy McGuine, Kristin Haraldsdottir, Kevin Biese, Leslie Goodavish, Bethany Stevens, Andrew M. Watson

**Affiliations:** Departments of Pediatrics, University of Wisconsin School of Medicine and Public Health, Madison, WI; Departments of Orthopedics and Rehabilitation, University of Wisconsin School of Medicine and Public Health, Madison, WI; The Department of Kinesiology, University of Wisconsin – Madison

**Keywords:** adolescent, infection, pediatric, SARS-CoV-2

## Abstract

**Introduction:** The purpose of this study was to describe the reported incidence of COVID-19 in Wisconsin high school athletes in September 2020, and to investigate the relationship of COVID-19 incidence with sport and face mask use.

**Methods:** Surveys were sent to athletic directors of all Wisconsin high schools regarding sports during September 2020. The association between reported case rates in athletes in each county and the county general population were evaluated with a weighted linear model. Multivariable negative binomial regression models evaluated the associations between COVID-19 incidence and sport type and face mask use by players, adjusting for the county COVID-19 incidence for each school.

**Results:** 207 schools that had reinitiated sport reported 270 COVID-19 cases among 30,074 players, for case and incidence rates of 809 cases per 100,000 players and 32.6 cases per 100,000 player-days, respectively. The case rates for athletes in each county were positively correlated with the case rates for the county’s general population (β =1.14±0.20, r=0.60, p<0.001). One hundred fifteen (55%) of cases were attributed to household contact, 85 (41%) to contact outside sport or school, 5 (2.4%) to school contact, and 1 (0.5%) to sport contact. No difference was identified between team and individual sports (incidence rate ratio (IRR)=1.03 [95% CI=0.49-2.2], p=0.93) or between non-contact and contact sports (IRR=0.53 [0.23-1.3], p=0.14), although the difference between outdoor and indoor sports approached statistical significance (IRR=0.52 [0.26-1.1], p=0.07). 84% of schools required face masks while playing. For those sports with >50 participating schools, there were no significant associations between COVID-19 incidence and face mask use in cross country (IRR=0.71 [0.2-2.2], p=0.52), football (IRR=1.6 [0.6-5.1], p=0.404), boys soccer (IRR=2.3 [0.5-17], p=0.31), or girls volleyball (IRR=1.4 [0.3-6.6], p=0.64).

**Conclusions:** Incidence of reported COVID-19 among athletes was related to background county incidence and most cases were attributed to household and community contact. Although not statistically significant, reported COVID-19 incidence may be lower in outdoor sports. Face mask use did not have a significant benefit, which may be due to relatively low rates of COVID-19 and the small number of schools that did not report using face masks.

## INTRODUCTION

The COVID-19 pandemic has caused unprecedented changes to the daily lives of people of all ages globally. Sports throughout the country have been shut down or altered in varying ways depending on the local county or state ordinances. In Wisconsin, youth sports were effectively canceled by the Safer At Home order in March 2020.^1^ With the order overturned by the Wisconsin Supreme Court in May 2020, counties throughout the state instituted their own restrictions.^2,3^ Youth sports were restarted in certain counties around the state during the summer of 2020, while others did not reinitiate.^4^

There has been a dearth of information regarding the risk of COVID-19 infection and sport participation. However, it is widely accepted that transmission of the SARS-CoV-2 virus that causes COVID-19 is related to direct exposure to respiratory droplets and airborne transmission.^5,6^ Longer periods of time in close proximity to infected individuals may increase transmission risk, and high intensity exercise may potentiate the spread of respiratory droplets, according to the United States Center for Disease Control and Prevention (CDC).^5^ National, state, and local regulations have all suggested an associated higher transmission risk with contact sports, indoor sports and team sport participation.^7^ Little direct evidence from sports exists to support this. Media reports of COVID-19 transmission within youth sports are a cause for concern, although it is often unclear whether the risk is due to sport participation or gatherings peripheral to or separate from the sports arena.^8,9^ A recent study in a soccer club from Washington state found physically distanced youth soccer training to be safe and did not contribute to COVID-19 spread among child and adolescent participants.^10^ Another preprint publication regarding club soccer players nationwide found no difference in reported COVID-19 incidence among athletes participating in contact versus non-contact soccer.^11^ Finally, a third preprint study of data collected from high school athletic directors nationwide found that, although COVID-19 incidence was higher among indoor sports, very few cases of COVID-19 were reportedly attributable to sport contact and the overwhelming majority were attributed to household and community contacts.^12^

Several recommendations to minimize COVID-19 risk in youth sports have been published since the onset of the pandemic by various academic organizations, public health agencies, and national sport governing bodies.^7,13,14,15,16^ However, because little data is directly available for sport contexts, these recommendations are largely based on inpatient COVID-19 data, case studies, and expert opinion. Risk mitigation recommendations vary widely and the debate around facemask use while playing sports continues.^17^ The CDC and American Academy of Pediatrics (AAP) recommend against the use of facemasks during play if they inhibit breathing, become wet, or become a choking hazard, but strongly recommend the use of facemasks any time while not in the act of playing a sport.^14,18^ Therefore, the purpose of this study is to describe the incidence of reported COVID-19 in high school sports in Wisconsin and to understand the associations between COVID-19 incidence and sport type as well as face mask use among athletes.

## METHODS

### Study Design

All procedures performed in this study were deemed exempt from by the Institutional Review Board of the University of Wisconsin-Madison. In collaboration with the Wisconsin Interscholastic Athletics Association (WIAA), surveys were distributed to all high school athletic directors on October 1, 2020. In addition to school name and location, athletic directors were asked whether they had restarted participation in sports since the initial COVID-19 restrictions in the spring of 2020. Those schools that reported reinitiating sports were asked to provide the specific sports and the date of restarting, number of athletes, number of practices and games, and number of COVID-19 cases among athletes within each sport, as well as the reported sources of infections (if known) during the month of September 2020. Schools were asked about their type of instruction during September (virtual or in-person) and whether they required the use of face masks for players while playing. Schools were included if they had any sport that had restarted participation during September 2020.

### Statistical Analysis

Data were initially evaluated using descriptive statistics, including estimates of central tendency (mean, median) and variability (standard deviation, interquartile range, range) for continuous variables, and counts and percentages for categorical variables. Reported COVID-19 case rates were expressed as the number of reported cases per 100,000 players (cases / total number of players * 100,000) overall and for each sport. Duration of participation for each sport at each school was determined as the difference in days between the date of restarting and October 1, 2020, and player-days was determined as the product of the number of participating players and duration. Reported COVID-19 incidence rates were expressed as the number of reported cases per 100,000 player-days (cases / total number of player-days * 100,000) overall and for each sport, with confidence intervals calculated using an exact method.

In addition, the number of cases, total population, case rate and incidence rate during September were determined for each county in which a respondent high school was located from publicly available online information from the local health authority. In order to determine whether background county COVID-19 case rates were associated with reported COVID-19 case rates among high school athletes, the total number of athletes and reported COVID-19 cases were aggregated by county. For those counties with >100 athletes, the relationship between COVID-19 case rates among high school athletes and the general population were evaluated with a linear model weighted for the total population of each county.

For those sports with data from 50 or more schools, the relative risk of each sport was evaluated using a mixed effects negative binomial regression model to predict the number of COVID-19 cases for each team with local incidence, instructional delivery type, and sport as fixed effects, the log of player-days as an offset, and school as a random effect, yielding an incidence rate ratio (IRR) with “Cheer / Dance” as the reference (since this represented the median unadjusted incidence rate). To evaluate the relationship between reported COVID-19 incidence and sport characteristics, a multivariable negative binomial regression model was developed to predict the number of cases, with local incidence, sport location (indoor, outdoor), sport contact (contact, non-contact), sport type (team, individual), and school instructional delivery type as covariates, and the log of player-days as an offset.

To evaluate the association between overall COVID-19 incidence and reported face mask use, incidence rates and 95% confidence intervals were calculated within each sport with greater than 50 reporting schools for those reporting face mask use or not. Separate multivariable negative binomial regression models were then developed to predict the number of cases, with local incidence, instructional delivery type, and face mask use (yes/no) as covariates, and the log of player-days as an offset. Coefficients from the models were exponentiated to represent IRRs for binary variables and Wald confidence intervals were generated. Significance level was determined *a priori* at the 0.05 level and all tests were 2-tailed. All statistical analyses were performed in R.

## RESULTS

Two hundred forty-four schools submitted complete survey responses, of which 207 had restarted a fall sport. These schools represented 30,074 student-athletes that had participated in 16,898 practices and 4,378 games. One hundred eighty-seven schools (90.3%) reported utilizing in-person instruction during September 2020. Among the schools that had restarted participation, 270 cases of COVID-19 were reported, yielding a case rate of 892 cases per 100,000 athletes and an incidence rate of 32.6 (95% CI = 28.9-36.8) cases per 100,000 player-days. From September 6 to October 4, 2020, 2390 cases of COVID-19 were reported among 14-17 year-olds in Wisconsin, for a case rate of 1067 cases per 100,000 people and an incidence of 38.1 cases per 100,000 person-days.^19^ Of the cases with a reported known source, 115 (55%) were attributed to household contact followed by community contact outside sport or school (85, 41%), school contact (5, 2.4%), sport contact (1, 0.5%) and other (3, 1.4%). For those sports with greater than 50 participating schools, the incidence rate ranged from 13.3 (Tennis – Girls) to 45.2 cases per 100,000 player-days (Football), as shown in Figure 1 (full data available in Supplemental Table 1).

**Figure 1.**
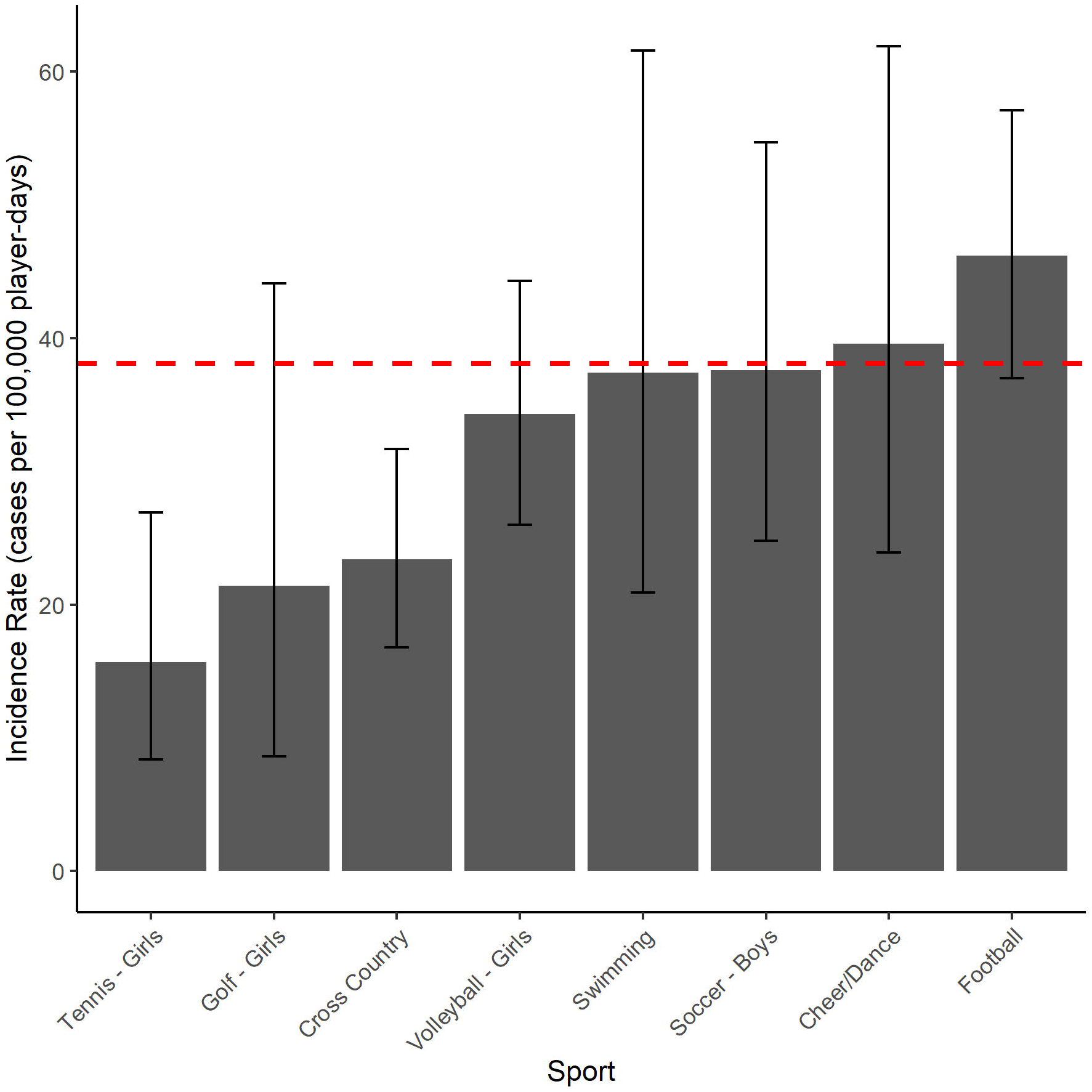
Unadjusted incidence rates of COVID-19 during September 2020 for various sports from Wisconsin high schools. Incidence rate is shown as reported cases per 100,000 player-days for those sports with greater than 50 schools reporting restarting. Red, dashed line represents the COVID-19 incidence rate among 14-17 year-olds in Wisconsin from 9/6 to 10/5/2020 (data extracted from https://www.dhs.wisconsin.gov/covid-19/cases.htm#youth on 2/5/2021).

When aggregated by county, the case rates for athletes in each county were significantly and positively correlated with the case rates for their respective county’s general population (β =1.14±0.20, r=0.60, p<0.001; see Figure 2). The IRRs for specific sports, adjusted for state COVID-19 incidence, instruction delivery type and school repeated measures are shown in Figure 3. The IRRs for school instructional delivery and sport characteristics are shown in Table 1. One hundred seventy-three schools (84%) reported face mask use by players while playing sports. Unadjusted incidence for teams with and without reported face mask use within each sport with greater than 50 respondent schools are shown in Figure 4. After adjusting for local county COVID-19 incidence and school instructional delivery, face mask use was not associated with a decreased COVID-19 incidence in football, girls’ volleyball, boys’ soccer or cross country (Table 2).

**Figure 2.**
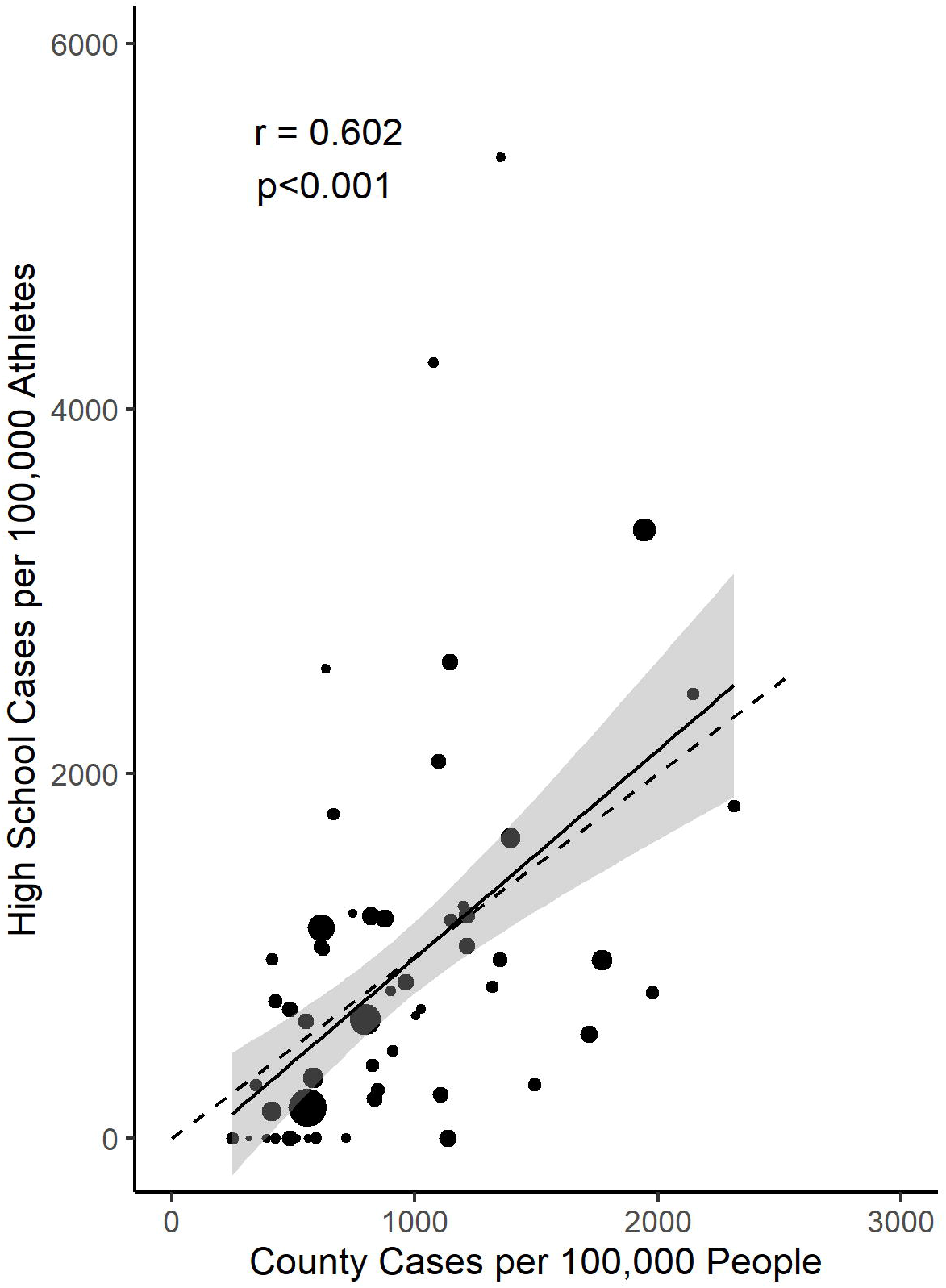
Reported COVID-19 case rates for Wisconsin high school athletes and the general population of their respective counties during September 2020. Size of points scaled to population of each county and dashed line represents a line of equality. Solid line and shaded area represent regression line and 95% confidence interval from linear model weighted for population of each county. r = correlation coefficient.

**Figure 3.**
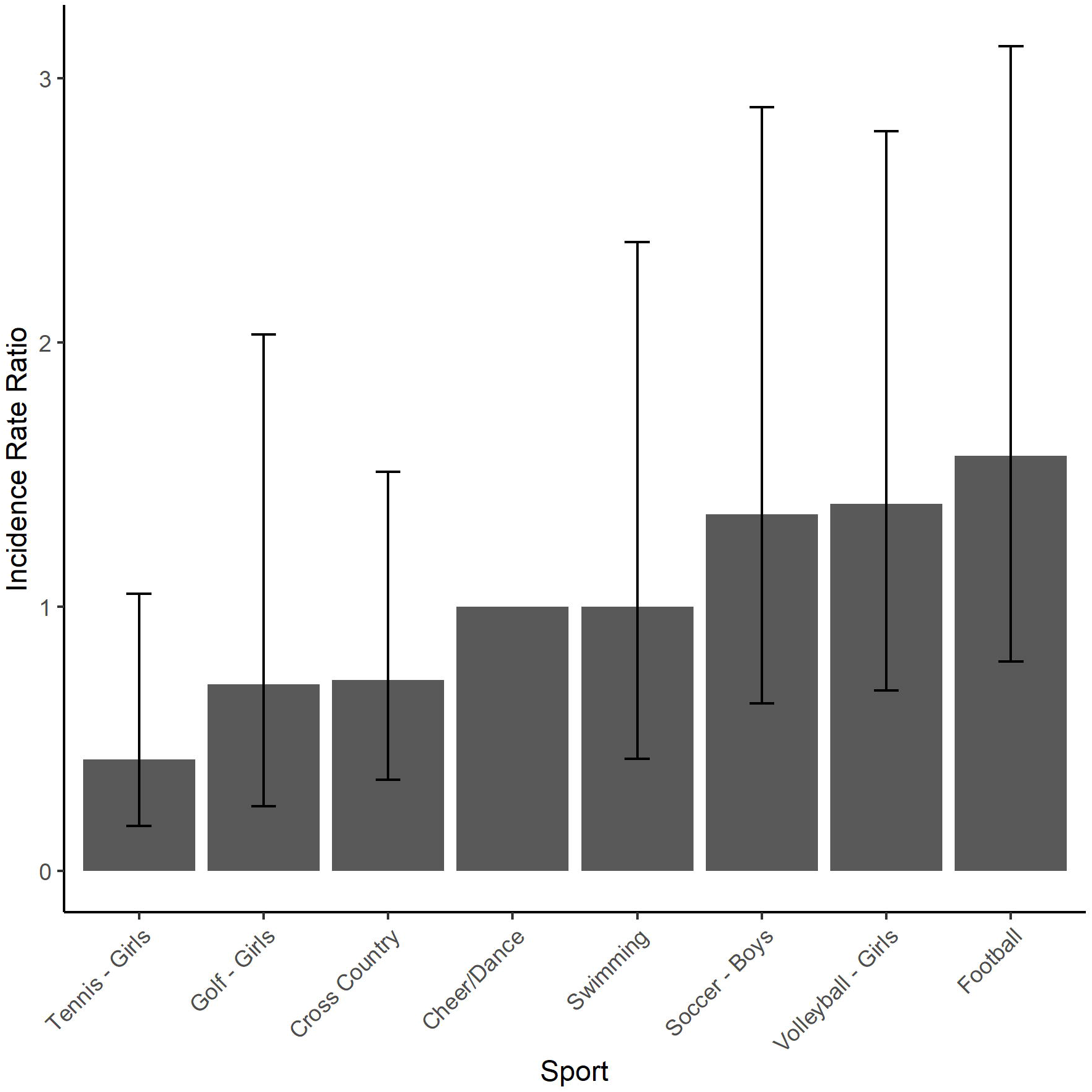
COVID-19 incidence rate ratios during September 2020 for Wisconsin high school sports, adjusted for local (state) COVID-19 incidence, instructional delivery type and repeated measures from the same school. Includes those sports with greater than 50 schools reporting participation, with Cheer/Dance as reference. *p<0.05.

**Table 1.** Incidence rate ratios for reported COVID-19 cases among Wisconsin high schools in fall 2020 by school instructional delivery and sport characteristics.

|  | IRR (95% CI) | p |
| --- | --- | --- |
| School instructional delivery (in-person) | 1.33 (0.74-2.4) | 0.34 |
| Outdoor | 0.52 (0.26-1.1) | 0.072 |
| Team | 1.03 (0.49-2.2) | 0.93 |
| Non-Contact | 0.53 (0.23-1.26) | 0.14 |

<sup>a</sup>Incidence rate ratios and Wald confidence intervals from a mixed effects negative binomial regression to predict COVID-19 cases with local incidence, instructional delivery type, and sport characteristics as fixed effects, school as a random effect and log(player-days) as an offset; CI = Confidence Interval; IRR=Incidence Rate Ratio.

**Figure 4.**
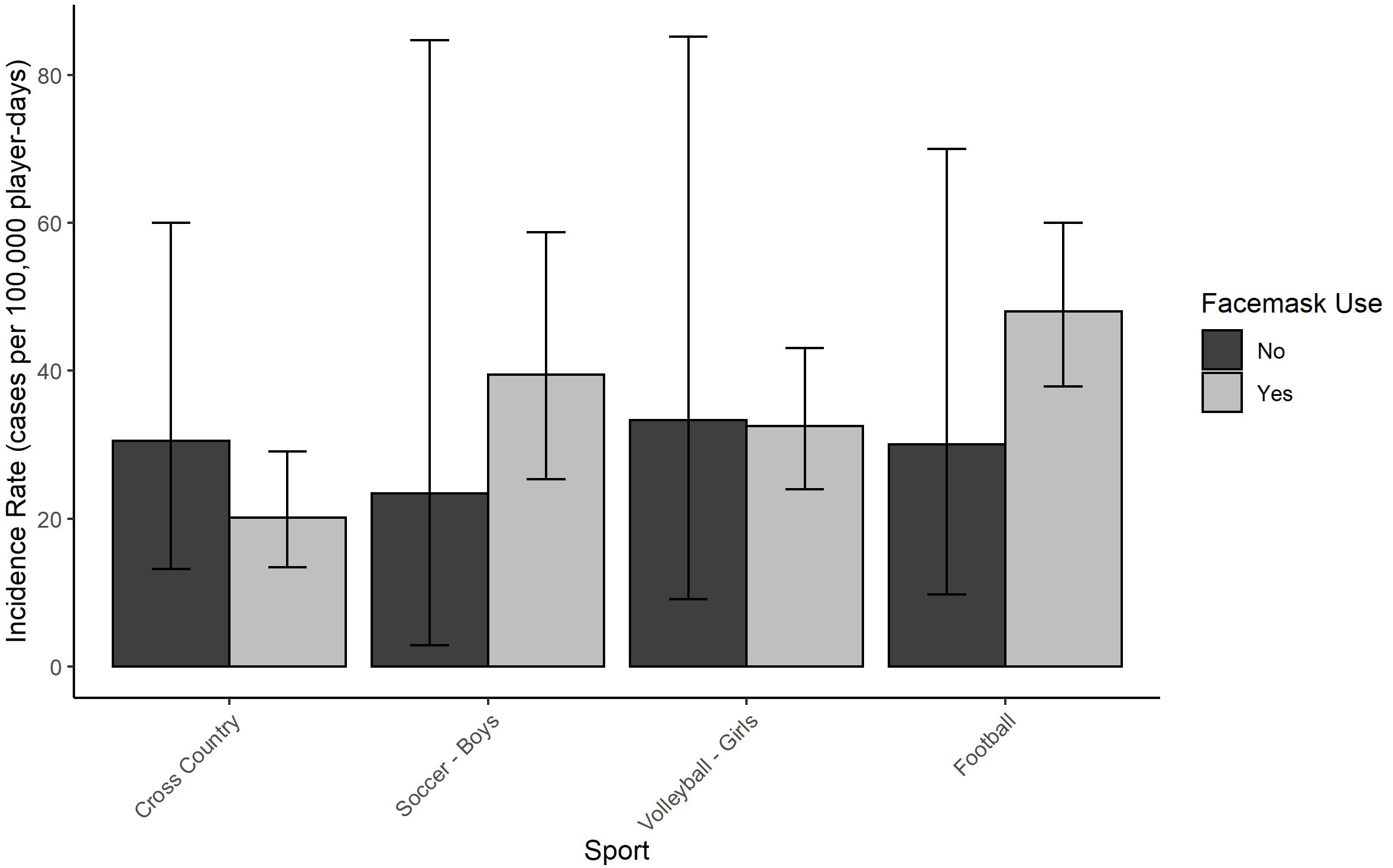
Unadjusted COVID-19 incidence rates reported among Wisconsin athletes in September 2020, comparing teams with or without reported face mask use, within each sport. Includes those sports with greater than 50 reporting schools.

**Table 2.** The association of reported face mask use with COVID-19 incidence within each sport among Wisconsin high school athletes during September 2020.^a^

|  | IRR (95% CI)<sup>a</sup> | p |
| --- | --- | --- |
| Cross Country | 0.706 (0.24-2.2) | 0.524 |
| Football | 1.58 (0.57-5.1) | 0.404 |
| Soccer - Boys | 2.34 (0.52-17) | 0.307 |
| Volleyball - Girls | 1.37 (0.3-6.6) | 0.641 |

<sup>a</sup>Incidence rate ratios and Wald confidence intervals from separate multivariable negative binomial regression models within each sport to predict COVID-19 cases with local incidence, instructional delivery type, and face mask use (yes/no) as fixed effects, and log(player-days) as an offset. CI = Confidence Interval; IRR=Incidence Rate Ratio.

## DISCUSSION

In this statewide survey study of high school athletes, we did not identify a statistically significant association between COVID-19 incidence and sport type after adjusting for local virus incidence and school instructional delivery. Girls’ tennis, girls’ golf and cross country reported the lowest adjusted incidence rates of COVID-19, while football reported the highest. Nonetheless, the confidence intervals around these estimates were wide, perhaps due to the relatively low incidence of COVID-19 in Wisconsin during this period. There were no independent, statistically significant differences in reported COVID-19 incidence between indoor versus outdoor sports, contact versus non-contact sports, or individual versus team sports. However, indoor and contact sport classification approached significance, suggesting that this relationship may become clearer with a larger sample during a period of higher COVID-19 incidence. In fact, a recent preprint publication among high school athletes nationwide demonstrated this relationship over a longer time period when background COVID-19 rates were higher throughout the country.^12^

We found that case rates of reported COVID-19 in high school athletes were significantly related to the case rates of their respective county’s general population, and very similar to the overall COVID-19 incidence rate among 14-17 year olds in Wisconsin during roughly the same time frame. In addition, 96% of the cases among athletes had a reported source of infection from household and community contacts, and only 1 (0.5%) from a known sport contact. This may suggest that the background, local COVID-19 incidence may have a greater effect on overall COVID-19 incidence among high school athletes than participation in a specific sport or type of sport.

Reported face mask use among the sports with the largest number of respondent schools (football, girls’ volleyball, boys’ soccer, and cross country) did not have a significant relationship with COVID-19 incidence. While this is similar to the findings among outdoor sports in a recent nationwide sample of high school athletes, it is in contrast to the finding from that study that facemask use was associated with decreased COVID-19 incidence among indoor sports.^12^ The overwhelming majority of the respondent schools in the present study (84%) reported facemask use while on the field or court. Together with the relatively low background COVID-19 incidence rates during September, this may have limited our ability to identify a true relationship between mask use and reported COVID-19 incidence.

Similarly, we did not find a relationship between COVID-19 incidence and type of instructional delivery (in-person, virtual). Of the cases with a reported known source, only 2.5% were attributed to a school contact. This is consistent with prior reports that schools have not been significant contributors to the spread of COVID-19.^20^ However, 90% of our respondent schools reported in-person instruction, making it difficult to fully evaluate the role of in-person school instruction in COVID-19 incidence among high school athletes. Nonetheless, we included school instruction type within our adjusted models in order to account for this as a potential confounder. Importantly, it should be recognized that this study cannot account for transmission or incidence of COVID-19 among attendees at high school sporting events beyond the participants. This represents an important potential contributor to community COVID-19 risk, and risk mitigation procedures should continue to be prioritized to protect both athletes and attendees.

### Limitations

This study has several limitations. We are unable to verify the information provided by athletic directors through a separate independent source. Local, county-level daily COVID-19 case data was often not available for adolescents or children, so our adjusted models could only account for the population-level background incidence from each county. Nonetheless, we found that reported case rates from our sample and the case rates from the county general populations were highly related. It is difficult to fully interpret comparisons of COVID-19 cases reported by athletic directors and those collected by public health agencies, but we have included public health data to add context for our findings and to adjust our incidence models. As mentioned above, the incidence of COVID-19 was relatively low during September 2020 in Wisconsin, and this may have limited our ability to detect statistically significant associations in some cases. Reported sources of infection were provided by the schools themselves and it is unknown whether these represent the results of formal contact tracing by local health authorities. Finally, this data represents information regarding athletes from a single state and may not be generalizable to other populations.

## Conclusions

After adjusting for local county COVID-19 incidence, no statistically significant differences in reported COVID-19 incidence were identified between sports or sport types among Wisconsin high school athletes during September 2020. Reported COVID-19 case rates among athletes were highly correlated with case rates for the general population for their respective counties, and incidence rates among athletes were very similar to those of 14-17 year olds in Wisconsin in general. Most cases among athletes were attributed to household and community contact with very few attributed to school or sport contacts. Further research is warranted to better define the risk factors for COVID-19 transmission during adolescent sport participation and the relative benefits of different risk mitigation strategies.

## Data Availability

Data will be made available by the study team on reasonable request.

## ACKNOWLEDGEMENTS

There are no funding sources to report for this study. Dr. Watson serves as the Chief Medical Advisor for the Elite Clubs National League and is supported by grants from the National Center for Advancing Translational Sciences (UL1TR002373; KL2TR002374). We are grateful for the resources and support of the UW Institute for Clinical and Translational Research. Dr. McGuine serves on the Sports Medicine Advisory Council for the National Federation of State High School Associations. There are no other relevant conflicts of interest to disclose.

