## Supplemental Table 1 for "Reported COVID-19 Incidence in Wisconsin High School Athletes During Fall 2020"

Supplemental Table 1. The number of participating schools, players, reported cases, player-days, COVID-19 case and incidence rates by sport among Wisconsin high schools in September 2020.

| Sport | Schools | Players | Practices | Games | Cases | Player-Days | Case Rate^a^ | Incidence Rate^b^ |
| --- | --- | --- | --- | --- | --- | --- | --- | --- |
| Basketball - Boys | 4 | 98 | 11 | 0 | 0 | 750 | 0 | 0 (0-492) |
| Basketball - Girls | 4 | 68 | 11 | 0 | 0 | 750 | 0 | 0 (0-492) |
| Cheer/Dance | 72 | 1337 | 1260 | 61 | 19 | 47945 | 1421 | 39.6 (23.9-61.9) |
| Cross Country | 185 | 4708 | 4076 | 811 | 41 | 175318 | 871 | 23.4 (16.8-31.7) |
| Field Hockey | 4 | 183 | 79 | 24 | 2 | 4283 | 1093 | 46.7 (5.65-169) |
| Football | 162 | 8228 | 2794 | 249 | 86 | 185961 | 1045 | 46.2 (37-57.1) |
| Golf - Girls | 68 | 831 | 1344 | 510 | 7 | 32694 | 842 | 21.4 (8.61-44.1) |
| Rugby | 1 | 60 | 11 | 0 | 0 | 1380 | 0 | 0 (0-267) |
| Soccer - Boys | 103 | 3147 | 1569 | 587 | 27 | 71774 | 858 | 37.6 (24.8-54.7) |
| Swimming | 62 | 1202 | 1323 | 278 | 15 | 40136 | 1248 | 37.4 (20.9-61.6) |
| Tennis - Girls | 78 | 2138 | 1510 | 665 | 13 | 82586 | 608 | 15.7 (8.38-26.9) |
| Volleyball - Boys | 19 | 580 | 278 | 133 | 2 | 13702 | 345 | 14.6 (1.77-52.7) |
| Volleyball - Girls | 179 | 7454 | 2626 | 1060 | 58 | 169251 | 778 | 34.3 (26-44.3) |
| Wrestling | 2 | 40 | 6 | 0 | 0 | 600 | 0 | 0 (0-615) |
| Total | 943 | 30074 | 16898 | 4378 | 270 | 827130 | 898 | 32.6 (28.9-36.8) |

^a^Case rates shown as cases per 100,000 players. ^b^Incidence rates shown as cases per 100,000 player-days (n, 95% confidence intervals).
